# The Effect of Exogenous Ketone Bodies on Cognition in Patients with Mild Cognitive Impairment, Alzheimer’s Disease and in Healthy Adults: A Systematic Review and Meta-Analysis

**DOI:** 10.1101/2025.09.17.25335999

**Authors:** Bruno Bonnechère, Elizabeth B. Stephens, Amy C. Boileau, Martin Ducker, Brianna J. Stubbs

## Abstract

**Importance:** Impaired cognitive function is a hallmark of neuropsychiatric disease, posing a significant challenge to patients, clinicians and healthcare systems. Emerging research on ketone bodies suggests they may function as an alternative fuel for the brain, potentially enhancing cognitive function through both metabolic and signaling pathways. An alternative to inducing ketosis by lowering dietary carbohydrate intake is consumption of exogenous ketones (EK).

**Objective:** It is unknown whether the existing literature collectively supports a beneficial effect of EK on cognitive function; this systematic review and metanalysis aims to aggregate available data and address this gap.

**Data Sources:** PubMed, Web of Science, and EMBASE databases were searched in October 2023 for key words and free words referring to ketone bodies, cognition, and health-related conditions.

**Study Selection:** Multiple reviewers selected 29 studies for inclusion in the analysis from the initial 1678 search results, which included randomized control studies of healthy participants and patients with neuropsychiatric conditions, using exogenous ketones as an intervention alongside a placebo, that included outcomes assessing cognitive function.

**Data Extraction and Synthesis:** A PRISMA model was used for abstracting data, and the PEDRo scale was used to assess study quality. Data was extracted and verified by independent investigators.

**Main Outcome:** Cognitive function measures.

**Results:** 29 studies (1,347 participants) were included, with 18 studies (875 participants) in the meta-analysis. Results indicate that EK administration has a modest but statistically significant positive effect on cognitive performance (SMD = 0.26, 95% CI: 0.11 – 0.40, p = 0.0007). Sub-group analyses showed no significant differences between study duration (acute vs. intermediate; p = 0.50), ketone form (mono-esters vs. medium-chain triglycerides; p = 0.06), population type (healthy vs. Alzheimer’s disease; p = 0.21), or the presence of acute cognitive stressors (p = 0. 25).

**Conclusions:** The findings suggest that EK could be a promising adjunctive strategy in dementia management, offering potential benefits even in patients who maintain sufficient carbohydrate intake. EK may provide psychiatrists with an innovative, non-invasive approach to supporting cognitive resilience in patients with neuropsychiatric disorders. Further clinical trials should refine the therapeutic application of EK and integrate them into comprehensive neuropsychiatric care protocols.

**Key points:** **Question:** Does the consumption of exogenous ketones improve cognitive function, and what are the variables that influence efficacy.

**Finding:** Exogenous ketones have a modest but significant effect on overall cognitive performance outcomes, with no clear effect of study duration or population, ketone form or dose, or the presence of a stressor.

**Meaning:** These findings strongly support further research to determine the ideal administration strategy for exogenous ketones to improve cognitive function.

## Introduction

Alzheimer’s Disease and related dementias (ADRD) are known to be a major public health problem for governments, particularly in those countries with ageing populations, such as the USA, UK and Japan [1]. In addition to the reduced quality of life and wellbeing for the patient, ADRD also imposes a cost on society through the significant financial burden of patient care [2]. Although pharmaceutical industries have tried to address these challenges by developing both symptomatic and disease-modifying treatments, any such treatments have to be considered in the context of possibly harmful side-effects and practicalities of long-term administration. For these reasons, there has been considerable interest in non-pharmacological strategies, such as ketogenic diets, for improving cognition in patients with mild cognitive impairment (MCI) and ADRD [3].

The canonical primary role of ketone bodies (or ketones) including β-hydroxybutyrate (BHB), acetoacetate (AcAc), and acetone, is to act as a metabolic substrate for brain function during development, and in settings of low carbohydrate availability. Indeed, classic experiments by Owen *et al* [4] found that ketones entering the brain via select, widely expressed mono-carboxylate transporters [5] can account for up to 60% of brain metabolic needs during prolonged starvation. Alongside their role as an alternative substrate, ketones have been hypothesized to have multiple non-energy, signaling effects in the brain, including increasing cerebral blood flow [6–9], modulating release of neurotransmitters [10, 11] and neurotrophins [12, 13], and altering proteostasis [9, 14, 15]. Taken together, there is a strong mechanistic rationale supporting functional benefit of strategies that increase the availability of ketones to the brain.

Ketones are produced endogenously, as a result of increased peripheral lipolysis and hepatic conversion of free fatty acids to ketone bodies [16]. Ketosis is typically defined as a blood BHB concentration of > 0.5 mM [17–20]. Endogenous ketone production increases as a result of dietary strategies that severely restrict carbohydrate intake, such as during voluntary or involuntary fasting [21] or with consumption of a low-carbohydrate, high-fat, ketogenic diet [17]. A healthy adult can produce up to ∼150 g of ketone bodies during a prolonged fast [22], reaching a physiological ketosis of 5 – 7 mM [17, 21]. In contrast, long-term consumption of a well formulated ketogenic diet results in a more modest ketosis of < 1 mM [17]. Whilst dietary strategies to augment ketosis have been popularized of late, there are perceived barriers to widespread implementation that include long-term poor adherence and concerns about the impact of a poorly formulated ketogenic diet on cardiovascular risk.

Exogenous ketones (EK) represent an alternative to existing dietary strategies and can increase circulating ketone concentrations to 0.5 – 5 mM in a rapid and dose-dependent manner, even when consumed alongside carbohydrate that would usually prevent endogenous ketosis [23–25]. There are several types of EK compounds that either directly contain ketone bodies (i.e., free BHB acid, BHB or AcAc mineral salts, esters of BHB or AcAc), or contain precursors that are readily metabolized into ketone bodies (i.e., ketogenic medium chain triglycerides (MCT), medium chain fatty acid esters, ketogenic alcohol (R)-1,3-butanediol). As a category, EK have no currently known safety concerns and, although some gastrointestinal symptoms can occur which may reduce long term compliance, these appear to be reduced by a gradual increase in daily dose [32, 44–47]. In the last decade, there has been a steady increase in the number of publications that have addressed the possible physical and cognitive effects of different EK compounds in healthy adults and those with neuropsychiatric disorders.

As a result of the known metabolic and signaling effects on ketone bodies within the brain, there has been a growing scientific and medical interest in understanding the impact of ketone bodies on cognitive function in those with neurological conditions and healthy individuals. Given the higher chance of detecting functional improvements when there is an existing deficit, and the prominent role of deficits in brain energy metabolism in neurodegenerative disease [26], it is perhaps unsurprising that the bulk of studies investigating the cognitive effect of ketosis have focused on disease populations. In a key recent systematic review, ketogenic diet and other ketogenic interventions were suggested to positively impact cognition in patients with Alzheimer’s Disease (AD) [27]. Furthermore, ketosis may have a positive effect on cognition even in healthy adults; a recent compelling triangulation of observational studies and mendelian randomization studies found that increased circulating BHB improved general cognitive function as well as delaying the risk of cognitive decline and AD [28]. However, to our knowledge no publications to date have performed a systematic review or meta-analysis to consolidate the evidence for the cognitive effects of EK alone in humans – with or without cognitive impairment.

To address this gap, we undertook this systematic review and metanalysis to evaluate the literature on the effect of EK on human cognition. Given the strong mechanistic evidence for ketones as a fuel and a signal in the brain, we aimed to determine whether EK had a beneficial effect on cognition. If a positive effect was found, we further planned to address if there was an impact of dose or EK compound, duration of intervention, as well as if any benefit was present in healthy volunteers, as well as patients with neuropsychiatric disorders.

## Methods

This systematic review was conducted in accordance with the Preferred Reporting Items for Systematic Reviews and Meta-Analyses (PRISMA) checklist [29] and was registered on PROSPERO (registration ID: CRD42023471727). The software program, Rayyan [30], was used during all stages of study selection and data extraction. For the present study, no ethics committee approval was necessary.

### Search Strategy

Records were searched on three databases (PubMed electronic database of the National Library of Medicine, the Web of Science database, and Embase) to identify eligible studies published before 31^st^ October 2023. MeSH terms and free words referring to ketone bodies (‘ketone bodies’ OR ‘ketosis’ OR ‘ketogenic’ OR ‘ β-hydroxybutyrate’ OF ‘BHB’ OR ‘beta-hydroxybutyrate’ OR ‘acetoacetate’ OR ‘ketone’ OR ‘exogenous ketone’ OR ‘ketone ester’ OR ‘ketone salt’ OR ‘medium chain triglyceride’ OR ‘MCT’), cognition ((“cognit*” OR “memory” OR “learning” OR “attent*” OR “intellect” OR “executive funct*” OR “recognit*” OR “IQ” OR “problem solving” OR “psychomotor speed” OR “mental flexib*” OR “choice react*” OR “emotional bias” OR “planning” OR “response inhibition”)), and health-related conditions (‘dementia’ OR ‘mild cognitive impairment’ OR ‘MCI’ OR Alzheimer’s Disease’ OR ‘AD’ OR ‘Parkinson’s Disease’ OR ‘PD’ OR ‘traumatic brain injury’ OR ‘TBI’ OR ‘concussion’ OR ‘CTE’ OR ‘stroke OR ‘multiple sclerosis’ OR ‘MS’) were used as keywords. References from selected papers and from other relevant articles were screened for potential additional studies in accordance with the snowball principle. Search was limited to journal articles published in English.

Two reviewers (ACB and BJS) independently screened titles and abstracts according to the inclusion/exclusion criteria. In the case of conflicts, both reviewers met to discuss their study selection decisions until they reached a mutual decision. Studies that were included based on title and abstract, advanced to full-text screening where the same process was followed (ACB and BJS review independent, resolved conflicts together). A reason for exclusion was provided during full-text screening. The final analyses were conducted using studies that advanced through both levels of screening.

### Eligibility criteria

A PICOs approach was used as inclusion and exclusion criteria:

- *Participants*: Healthy participants and patients with neuropsychiatric conditions.
- *Intervention*: The intervention being studied was exogenous ketone products. This term encompasses any product that is administered with the aim of increasing blood ketone concentrations. These may include, but not be limited to ketone esters, ketone monoester, ketone di-ester, butanediol, ketone salt, medium chain triglycerides, tri-octanoate, coconut oil, ketone infusion.
- *Control*: Placebo
- *Outcomes*: Outcomes encompassed assessments of one or more cognitive functions conducted both prior to and following ketone body supplementation. These outcomes included a comprehensive evaluation of global cognition, as well as various subdomains of cognitive function, which include verbal memory, nonverbal memory, working memory, processing speed, attention, language proficiency, visuospatial skills, and executive functions. The evaluation of cognitive functions must have been conducted by qualified healthcare professionals such as psychologists, medical doctors, or neuropsychologists
- *Study design*: RCTs

A flow diagram of the study selection with the screened articles and the selection process is presented in **Figure 1**.

### Data extraction

Data were extracted independently by a single reviewer (EBS) and confirmed by a second reviewer (BJS) to verify correct data extraction. The following information was extracted from the included studies: characteristics of the patients (age, sex ratio, general information about conditions/disease), characteristics of the intervention (molecule, dose, duration), type of control and outcomes measurements. Means and error (as standard error, standard deviation, or 95% CI) for cognitive function scores were extracted either by directly extracting from tables/text or by using the data extraction tool, WebPlotDigitizer (WebPlotDigitizer, Pacifica, CA, USA), when data were only reported in figures. Participant population, number of participants, and any co-interventions were also extracted.

### Quality assessment

The PEDro scale, which is deemed a valid and reliable tool for assessing RCTs, was used for methodological quality assessment. RCTs’ quality was blindly judged by two different reviewers (BJS and EBS) to minimize potential bias. To ensure the rigor of this process, the final decision about each RCT quality was made by reaching a consensus. In case of discordance, a third collaborator (BB) was consulted to provide expert input. The RCTs were classified into distinct categories based on their quality: low quality (scores falling within the range of 0 to 3 out of 10; moderate quality (scores spanning 4 to 6 out of 10) and high quality (RCTs achieving scores from 7 to 10 out of 10). This methodical approach allowed for a comprehensive evaluation of the RCTs, thus facilitating an objective assessment of their respective quality levels.

### Statistical analysis

For studies assessing the efficacy of ketone bodies and displaying complete results of the pre and post-tests, we performed a meta-analysis. The measure of treatment effect was the standardized mean difference effect size (standardized mean difference [SMD]), defined as the between-group difference in mean values divided by the pooled SD computed using the Hedge’s g method. If different tests were used to assess the cognitive in the same study, the different results were pooled to have one unique SMD as recommended by Cochrane’s group. A positive SMD implies an increased cognitive function in comparison with the control group. We calculated the variance estimate tau² as a measure of between-trial heterogeneity. We prespecified a tau² of 0.0 to represent no heterogeneity, 0.0–0.2 to represent low heterogeneity, 0.2–0.4 to represent moderate heterogeneity, and above 0.4 to represent high heterogeneity between trials. To deal with high or moderate heterogeneity we used random-effect models and presented forest plots for the different comparators. We checked for publication bias using funnel plot [15] and Egger’s test for the intercept was applied to check the asymmetry [16] and performed sensitivity analysis to detect potential outliers. Meta-regression was performed to assess a possible effect of the dose, the duration (long term = > 13 days; immediate = single administration and same-day measurement) of the intervention and the type of supplementation on the results.

The statistics were conducted in RStudio (version 2023.06.2) with R version 4.4.2. and the significance level set at p < 0.05.

## Results

### Search results

Twenty-nine studies have been finally included in the systematic review. The PRISMA flowchart of the study selection is presented in **Figure 1**.

### Characteristics of the participants

**Table 1** provides a comprehensive overview of the 29 included studies, detailing their respective characteristics. In total 1347 participants were included in this review, amongst them 1288 (95%) completed the full protocol. The complete characteristics of the participants and interventions are presented in **Table 1**.

**Table 1:**
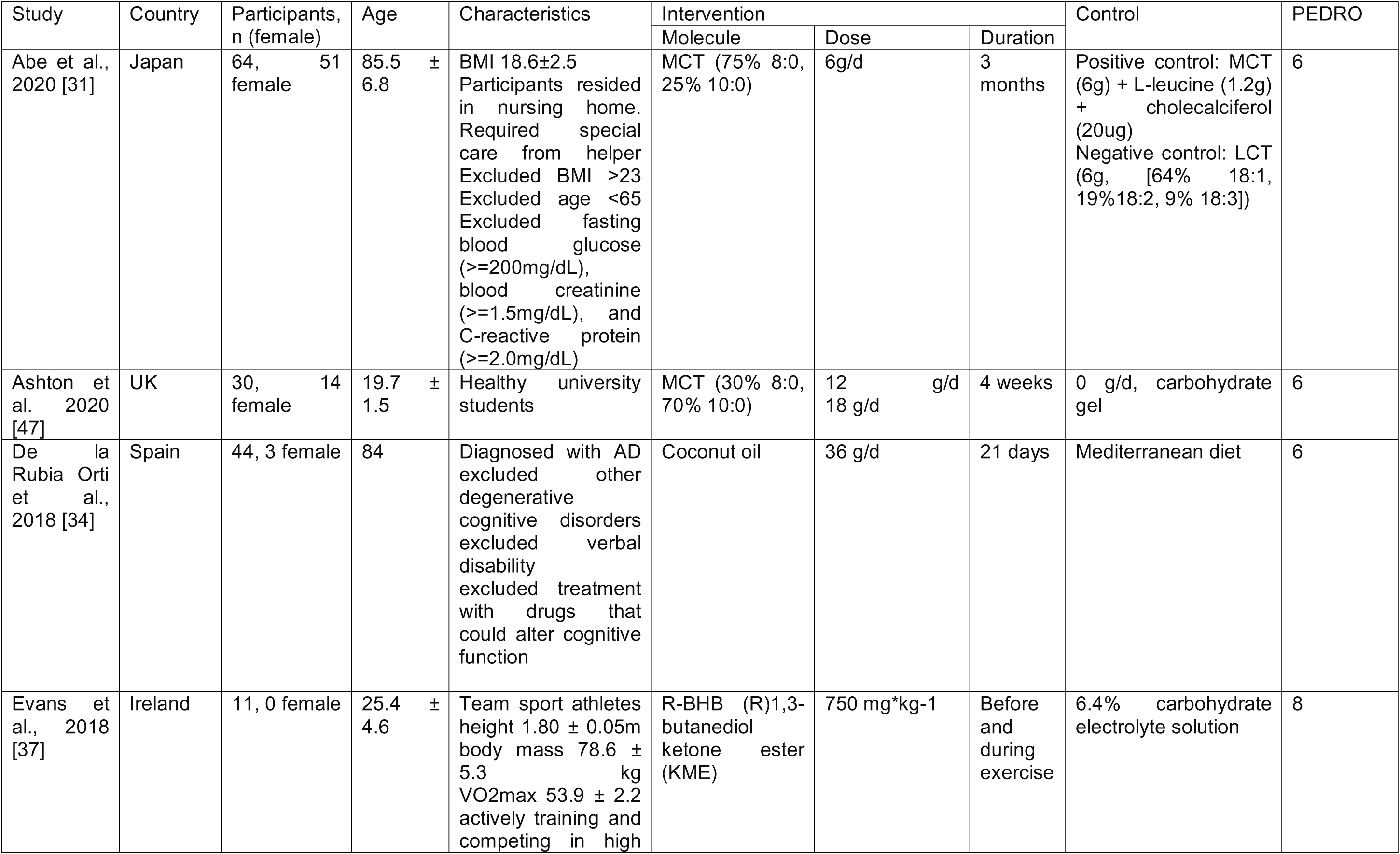
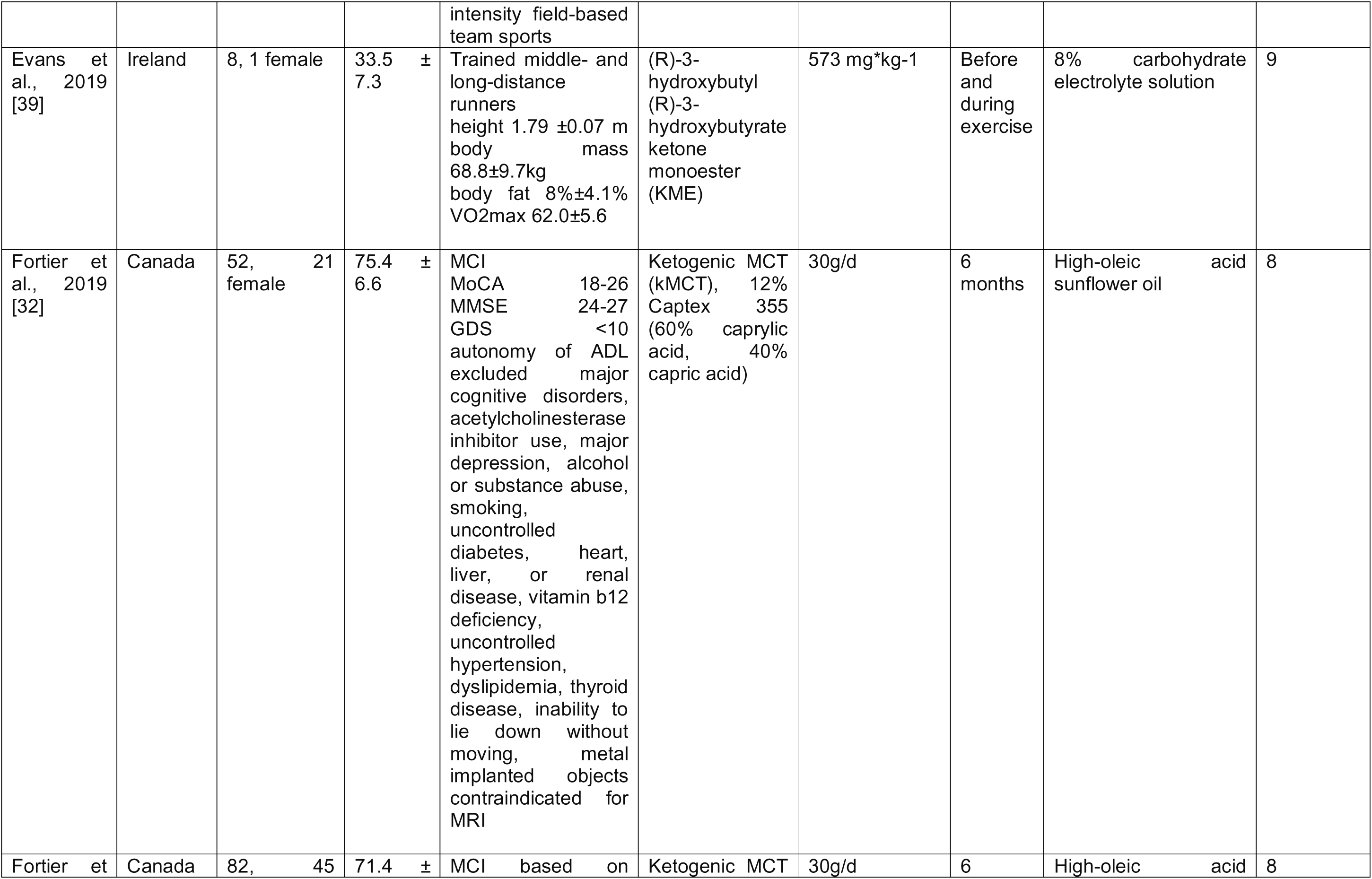
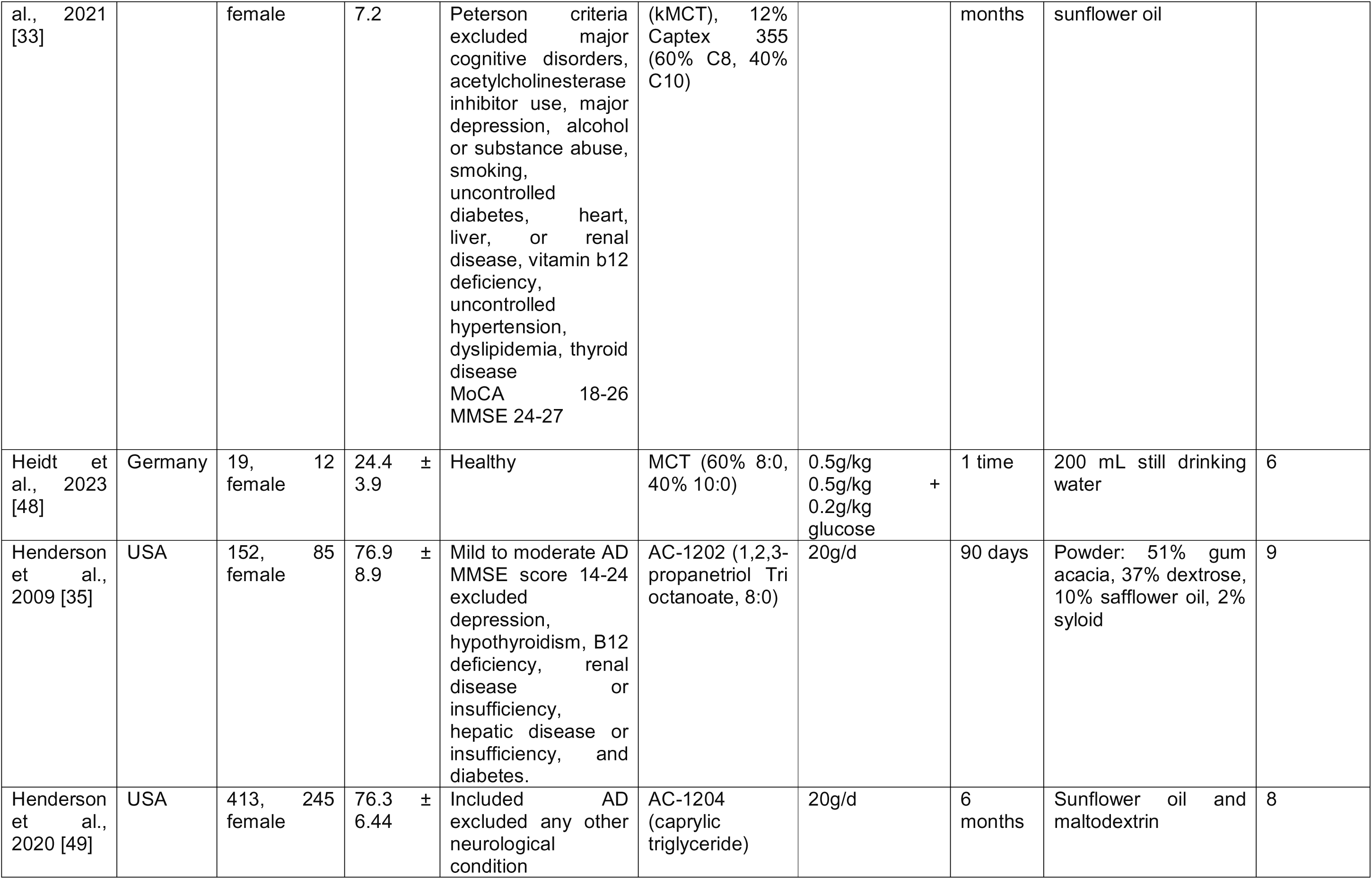
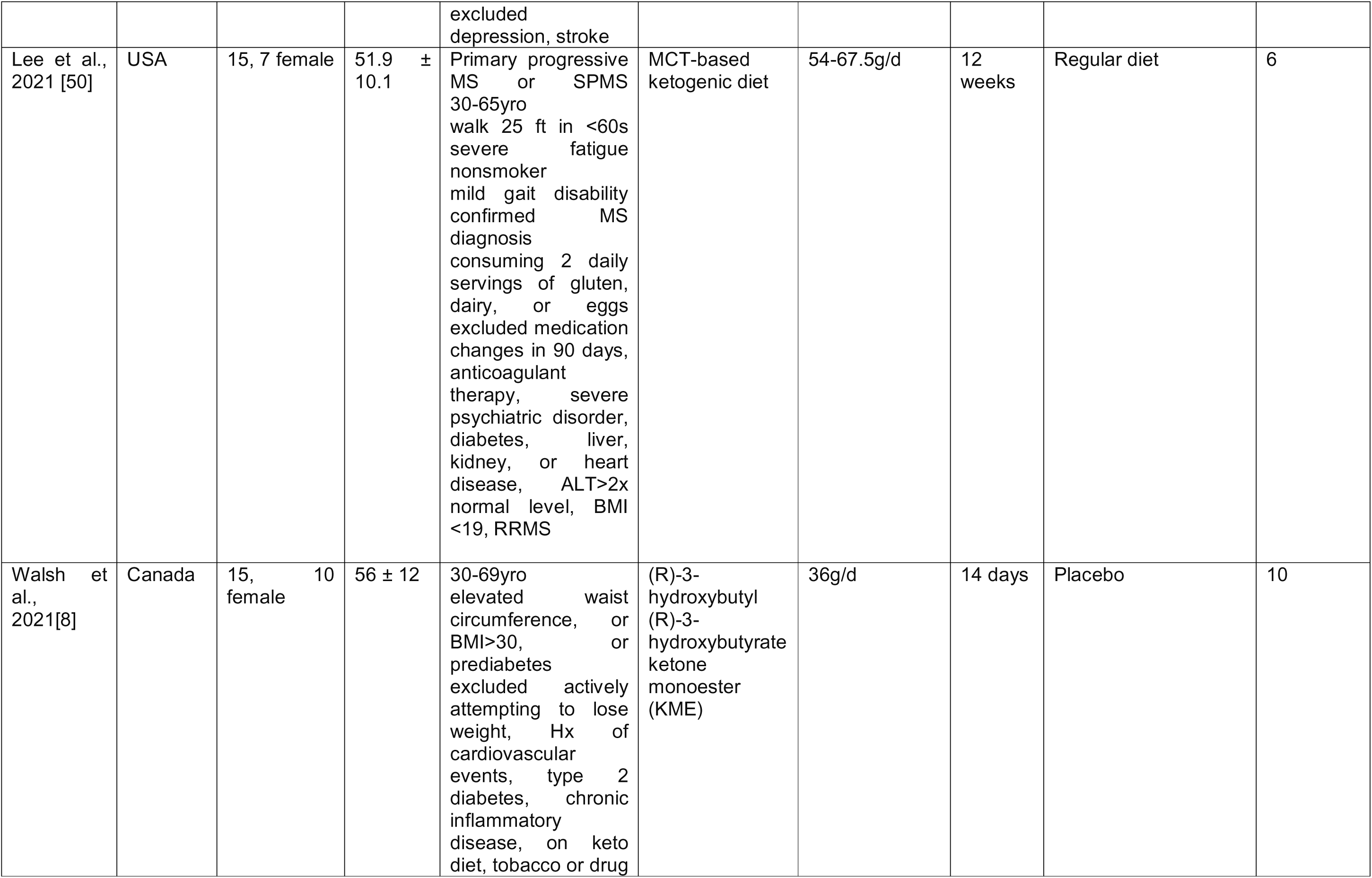
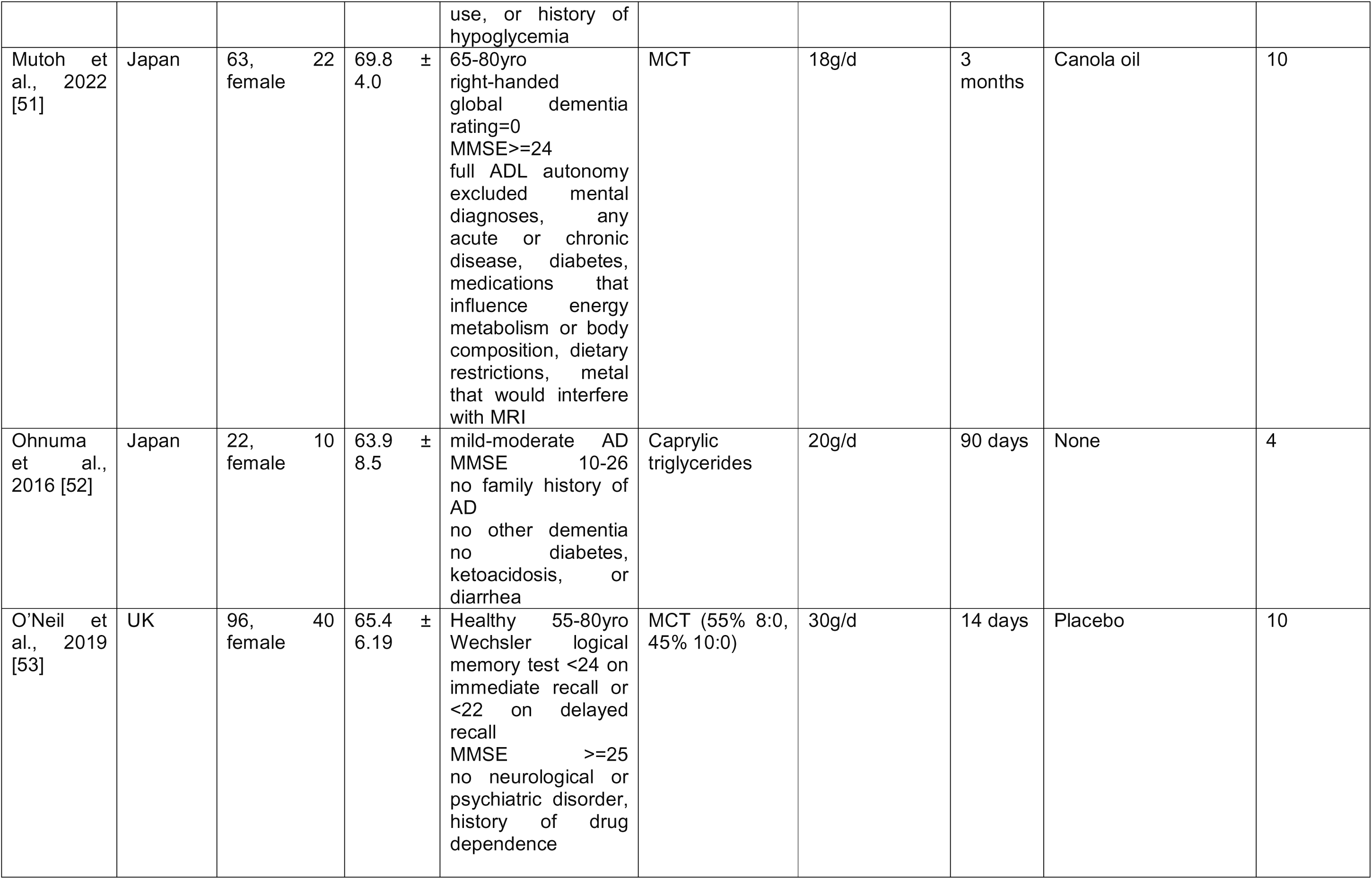
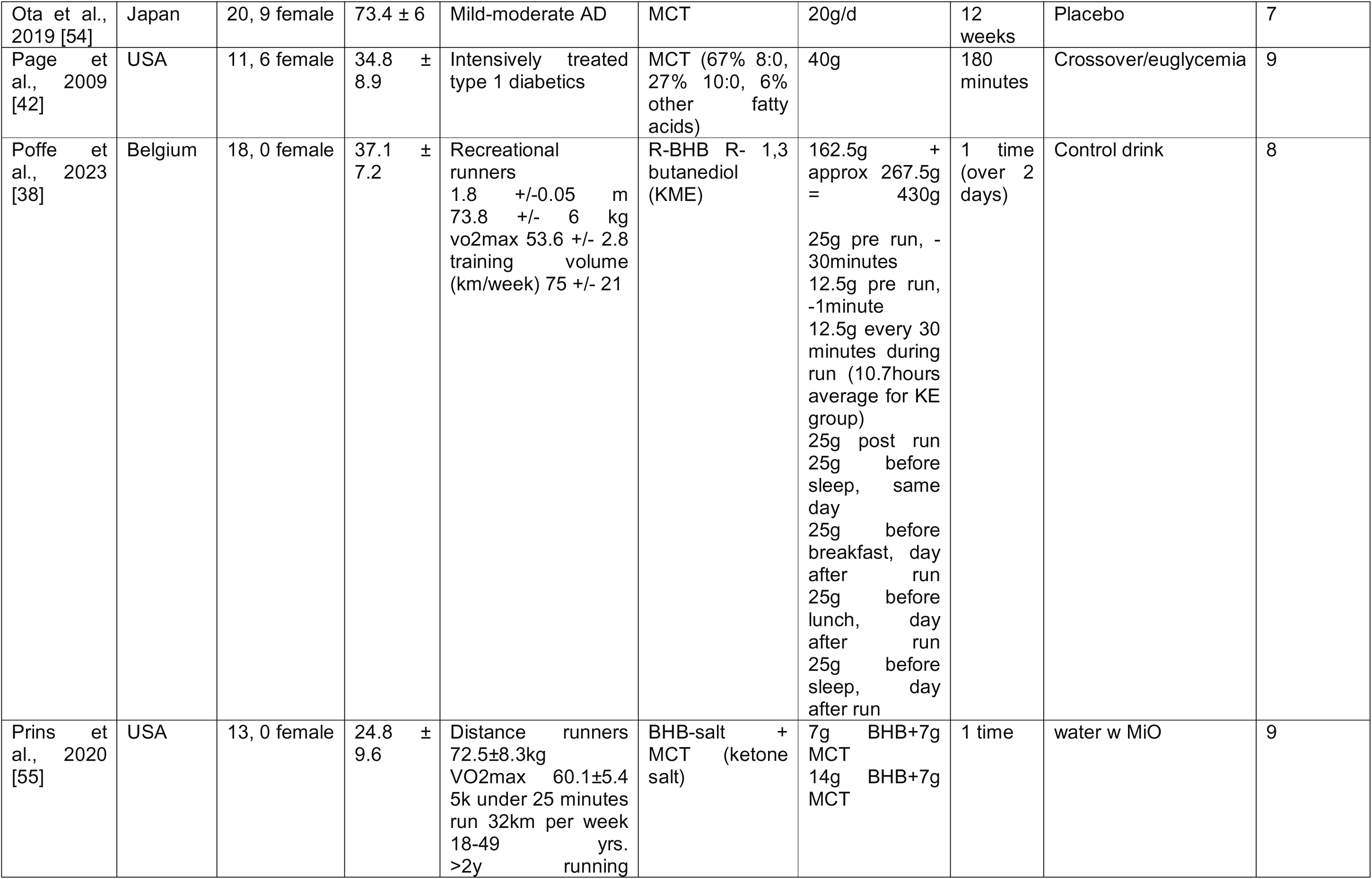
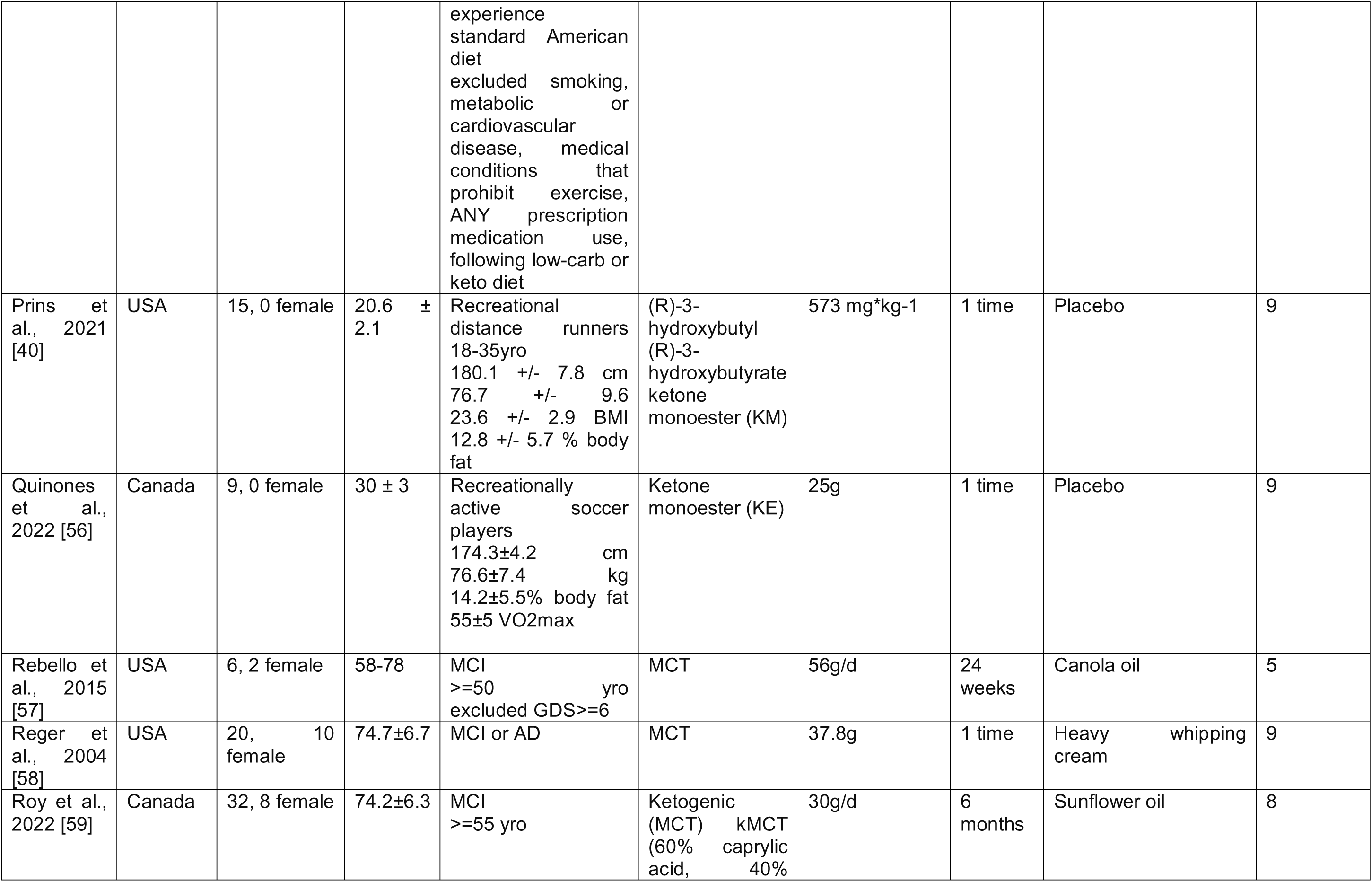
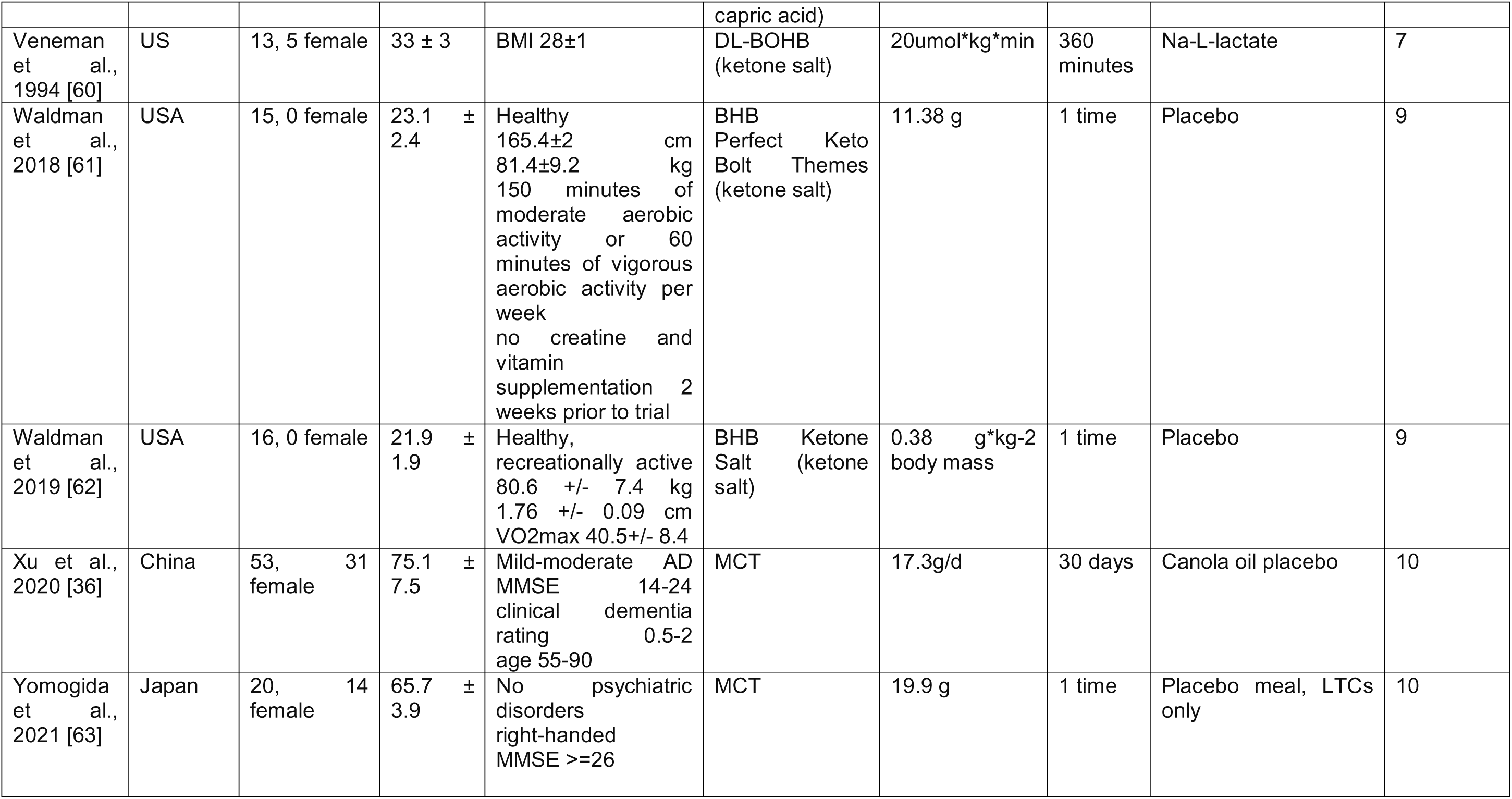
Characteristics of studies included in the systematic review and meta-analysis.

Concerning the quality of the individual studies we found an overall PEDRO score of 8 (1.6) out of 10 indicating high quality studies, with only seven studies being judged as moderate quality. Individual results are presented in **Table 1** and summarized in **Figure 2**.

## Systematic review

### Characteristics of the intervention

The studies reviewed encompass a wide range of geographic locations, with the largest number of participants coming from the United States, which contributed 664 participants across 9 studies. Japan followed with 189 participants across 5 studies, while Canada included 190 participants in 5 studies. Other countries with fewer participants included the United Kingdom (126 participants across 2 studies), Ireland (19 participants across 2 studies), Spain (44 participants in 1 study), Germany (19 participants in 1 study), Belgium (18 participants in 1 study), and China (53 participants in 1 study).

The types of supplementations presented high heterogeneity, with lipid-based precursors being the most common. MCTs were used in 1,038 participants across 18 studies, with dosages that ranged from 6 g/day to 56 g/day, with common formulations including caprylic acid (C8:0) and capric acid (C10:0). Coconut oil was used in a single study with 44 participants, administered at a dosage of 36 g/day, in the context of AD. AC-1202 and AC-1204 (proprietary, purified caprylic triglycerides), were used in studies involving 565 participants across 2 studies in the USA. Non-lipid-based ketone supplements included BHB salts, which were used in 44 participants across 2 studies, primarily in athletic performance contexts, and ketone monoester (KME), involving 172 participants across 6 studies. The dosages for the KME interventions varied, typically ranging from 573 mg/kg to 162.5 g total, and were predominantly used in studies focusing on athletic performance and cognitive function under stress.

The duration of the interventions also varied widely. The intervention duration ranged from single-dose studies to longer studies lasting up to 6 months, with the most common durations being between 1.5 to 3 months.

The most common outcomes evaluated across these studies focused on cognitive function and physical performance. Cognitive function was assessed using various tools, with the Mini-Mental State Examination (MMSE) and Alzheimer’s Disease Assessment Scale-Cognitive Subscale (ADAS-Cog) being the most frequently used. Neuropsychological tests such as the Trail Making Test and Stroop Test were also commonly employed. These assessments were applied across diverse populations, including healthy adults, individuals with MCI, and patients with AD. Physical performance outcomes were primarily measured in athletes and healthy adults, focusing on endurance performance (e.g., time trials), muscle strength (e.g., grip strength, knee extension time), and reaction times. Additionally, biomarkers like plasma BHB levels were often measured to confirm ketosis, alongside glucose and lactate concentrations, particularly in studies involving KME.

### Clinical efficacy

Our interest focused on the clinical effects of ketone supplementation on cognitive performance. The relevant outcomes and main results of the included studies are presented in **Table 2**.

**Table 2:** Description of participants’ outcomes and main tests of the included studies.

| Study | Summary<br>(Duration, type,<br>Participant health<br>status, stressor) | Outcome and tests | Main results | Conclusion |
| --- | --- | --- | --- | --- |
| Abe et al., 2020<br>[31] | Long-term, MCT,<br>healthy, no<br>stressor. | • **Primary Tests:** 10-second leg open/close test, MMSE.<br>• **Secondary Tests:** Upper-arm circumference, bilateral calf circumference, triceps skinfold thickness, mid-upper-arm muscle area, hand-grip strength, knee extension time, walking speed, peak expiratory flow, RSST (swallowing function), FIM (ADL). | MCT supplementation increased total MMSE score by 3.5 points from BL to 3-mo intervention | Supplementation with 6g MCT may improve cognition in frail older adults |
| Ashton et al.<br>2020 [47] | Long-term, MCT,<br>healthy, no<br>stressor | • **Primary Tests:** Trail Making Test, Digit Span Test (forward/backward), Spatial Span Test, Covert Shift of Attention, Rapid Visual Information Processing. | After 2-3 weeks of MCT supplementation, performance improved in cognitive tasks (trail making a/b, digit span forwards/backwards). there were minimal differences between the different doses | MCT ingestion improved cognitive performance after 2-3 weeks, minimal difference between 12g/d and 18g/d |
| De la Rubia Orti et al., 2018 [34] | Long-term,<br>coconut oil, AD,<br>no stressor | • **Primary Tests:** Benton's Temporal Orientation Test, Clock Drawing, Categorical Verbal Fluency, Free and Cued Selective Reminding Test. | After intervention, episodic, temporal orientation, and semantic memory improved. effect is more evident in women with mild-moderate state AD | The coconut oil enriched mediterranean diet improves cognitive function in patients with AD |
| Evans et al.,<br>2018 [37] | Immediate, KE,<br>healthy, exercise | • **Primary Tests:** Reaction Time Test.<br>• **Secondary Tests:** VO2max, | The KE resulted in increased plasma BHB concentrations. Plasma glucose and lactate | KE ingestion attenuated a decline in executive function after exhausting exercise, |
|  |  | Loughborough Intermittent Shuttle Test (LIST) | concentrations were lower in the KE group. HR, RPE, and 15m sprint times did not differ. Run time to exhaustion was not different. Incorrect responses in a multitasking test increased in the PLA group but not KE | suggesting a cognitive benefit. |
| Evans et al., 2019 [39] | Immediate, KE, healthy, exercise | • **Primary Tests:** 10 km time trial.<br>• **Secondary Tests:** Cognitive performance tests, oxygen consumption, running economy, respiratory exchange ratio (RER), HR, RPE, plasma BHB, glucose, lactate concentrations | Following KME ingestion, plasma BHB concentrations increased, but glucose and lactate concentrations were similar. No other measures differed. | KME ingestion by endurance-trained athletes elevated plasma BHB concentrations but did not improve 10km running time trials or cognitive performance. |
| Fortier et al., 2019 [32] | Long-term, MCT, AD, no stressor | • **Primary Tests:** Positron Emission Tomography (PET) for brain metabolism.<br>• **Secondary Tests:** RL/RI16, Brief Visual Memory Test-Revised, Trail Making Test, Stroop Color and Word Interference, Verbal Fluency, Digit Symbol Substitution, Boston Naming Test. | Brain ketone metabolism increased by 230% for kMCT. kMCT improved measures of episodic memory, language, executive function, and processing speed | This dose and duration improve several cognitive outcomes in MCI |
| Fortier et al., 2021 [33] | Long-term, MCT, AD, no stressor | • **Primary Tests:** RL/RI16, Verbal Fluency, Boston Naming Test, Trail Making Test. | Free and cued recall, verbal fluency, Boston naming test, and trail making tests improved with kMCT. Some cognitive outcomes correlated positively with plasma ketones. | This drink improved cognitive outcomes in MCI |
| Heidt et al., 2023 [48] | Immediate, MCT, healthy, no stressor | • **Primary Tests:** Wechsler Adult Intelligence Scale-IV (WAIS-IV) Verbal Comprehension Index (VCI), Working Memory Index (WMI).<br>• **Secondary Tests:** Satiety and tolerability questionnaires, plasma BHB levels. | Plasma BHB increased after ingestion of MCT oil, and a delayed increase with MCT + glucose. MCT oil plus glucose showed improved scores for the cognitive tests | For MCT + glucose consumption, there were fewer side effects and positive effects on cognitive ability, and similar trends were also observed for MCT alone |
| Henderson et al., 2009 [35] | Long-term, MCT, AD, no stressor | • **Primary Tests:** ADAS-Cog, ADCS-CGIC.<br>• **Secondary Tests:** Serum BHB levels. | Molecule elevated serum BHB. AC-1202 group had significantly different mean change in ADAS-Cog | AC-1202 rapidly elevated serum ketone bodies in AD patients and resulted in significant differences in ADAS-Cog scores compared to the Placebo. Effects were most notable in APOE4(-) subjects who were dosage compliant. |
| Henderson et al., 2020 [49] | Long-term, MCT, AD, no stressor | • **Primary Tests:** ADAS-Cog11.<br>• **Secondary Tests:** ADCS-ADL, Clock Drawing, Resource Utilization in Dementia (RUD-Lite), QoL in Alzheimer's Disease (QoL-AD), MMSE. | AC-1204 was tolerated. Mean changes in ADAS-Cog11 for placebo was 0.0, AC-1204 was 0.6 | AC-1204 did not improve cognition or functional abilities in subjects with mild-moderate AD |
| Lee et al., 2021 [50] | Long-term, MCT, healthy, no stressor | • **Primary Tests:** Modified Fatigue Impact Scale (FIS), Expanded Disability Status Scale (EDSS).<br>• **Secondary Tests:** MSQoL, Multiple Sclerosis Functional Composite (MSFC). | BHB indicated nutritional ketosis in keto group, in paleo and control, no change. paleo group had reduction in fatigue scores and maintained cognitive function scores. keto group had reduction in fasting glucose in insulin. | Consuming the MCT-based ketogenic diet achieved nutritional ketosis, but it was not associated with clinical improvement, whereas the paleo diet saw clinical improvements. |
| Walsh et al., | Long-term, KE, | • **Primary Tests:** Dual-task | DSST performance improved | BOHB supplementation |
| 2021[8] | healthy, no stressor | performance test, Stroop Task, Task Switching.<br>• **Secondary Tests:** Cerebral blood flow, BDNF levels, cerebrovascular conductance. | and was associated with improvements in cerebrovascular outcomes. | improved cognition in adults with obesity |
| Mutoh et al., 2022 [51] | Long-term, MCT, healthy, no stressor | • **Primary Tests:** Brain PET scan, Resting-state fMRI, MMSE.<br>• **Secondary Tests:** Wechsler Memory Scale-Revised (WMS-R) Logical Memory (LM) test, Trail Making Test, Digit Span, Digit Symbol Substitution Test (DSST), physical function, and anthropometric measures. | MCT group had better balance ability, no cognitive or other gait parameter change. MCT suppressed glucose metabolism | 3-month MCT supplementation improves walking balance |
| Ohnuma et al., 2016 [52] | Long-term, MCT, AD, no stressor | • **Primary Tests:** Alzheimer's Disease Assessment Scale-Cognitive Subscale (ADAS-Cog), MMSE. | Intervention did not improve cognitive function. Some ApoE4-negative patients with baseline MMSE $\geq 14$ showed improvements | Might be effective for AD patients with MMSE $\geq 14$ |
| O'Neil et al., 2019 [53] | Long-term, MCT, healthy, no stressor | • **Primary Tests:** Cambridge Neuropsychological Test Automated Battery (CANTAB), Source Memory Task. | Increased plasma BHB concentrations No significant improvements in cognitive function or memory related neuronal activity observed | Increasing plasma BHB levels with this intervention had no effects on cognitive function. |
| Ota et al., 2019 [54] | Immediate, MCT, AD, no stressor | • **Primary Tests:** Wechsler Memory Scale-Revised (WMS-R), WAIS-III, Trail Making Test, Stroop Test. | No significant difference between the one-time MCT dose and placebo for cognitive function.<br>In 12 week, trial, improvement in Digit symbol coding and immediate logical memory. | The chronic consumption of this formula may have positive effects on verbal memory and processing speed in patients with AD |
| Page et al., 2009 | Immediate, MCT, | • **Primary Tests:** Immediate | Hypoglycemia impaired | MCT ingestion improves |
| [42] | healthy, hypoglycemia | Verbal Memory, Delayed Verbal Memory, Verbal Memory Recognition, Digit Span Backward, Letter/Number Sequencing, Digit Symbol Coding, Map Search (1 min and 2 min), Telephone Search. | cognitive function. MCT ingestion reversed these effects. MCT increased BHB levels. | cognition in response to hypoglycemia in T1D. |
| Poffe et al., 2023 [38] | Immediate, KE, healthy, exercise | Run until exhaustion, or until 100km ran<br>• **Primary Tests:** Reaction Time Test, Rapid Visual Information Processing, Spatial Working Memory, Cambridge Gambling Task. | In control, RUN increased visual reaction time and movement execution time, however KE negated this effect | KE enhances mental alertness in ultra-endurance exercise |
| Prins et al., 2020 [55] | Immediate, BHB salt + MCT, healthy, exercise | • **Primary Tests:** 5k time trial.<br>• **Secondary Tests:** Automated Neuropsychological Assessment Metric (ANAM). | Increased BHB, exercise performance was unaltered, however there were responders and non-responders. larger dose augmented cognitive function in pre-exercise conditions, and exercise increased cognitive performance for smaller dose and placebo. | This formulation had a dosing effect on cognitive performance, but did not influence exercise performance. |
| Prins et al., 2021 [40] | Immediate, KE, healthy, exercise | Voluntary hypoventilation with exercise<br>**Primary Tests:** Automated Neuropsychological Assessment Metric (ANAM). | KE increased BHB, reduced blood glucose, no changes in lactate production, alterations to pH, bicarbonate, total carbon dioxide, increased respiratory exchange ratio, and blood carbon dioxide was significantly lower immediately post voluntary hypoventilation | No difference between KE and PLA in cognitive decline resulting from voluntary hypoventilation plus exercise. |
|  |  |  | + exercise. The protocol decreased cognitive performance similarly in both conditions. |  |
| Quinones et al., 2022 [56] | Immediate, KE, healthy, exercise | • **Primary Tests:** Stroop Test, Choice Reaction Task. | KME had a reduced performance decrease compared to placebo. No other cognitive function differences see | KME supplementation attenuated decreases in the choice reasoning task during repeated, high intensity, intermittent exercise |
| Rebello et al., 2015 [57] | Long-term, MCT, MCI/AD, no stressor | • **Primary Tests:** ADAS-Cog. | MCT ingestion increased serum ketones and improved memory | Consumption of 56g/d of MCT for 24 weeks increases serum ketone concentration and may modulate cognitive function |
| Reger et al., 2004 [58] | Immediate, MCT, MCI/AD, no stressor | • **Primary Tests:** ADAS-Cog, MMSE, Stroop Test, Paragraph Recall. | MCT treatment facilitated higher performance on ADAS-Cog for E4- subjects | Pts with APOE4- patients may benefit from MCTs |
| Roy et al., 2022 [59] | Long-term, MCT, MCI, no stressor | • **Primary Tests:** Trail Making Test (TMT), Stroop Test, DSST, MRI, PET scan. | kMCT was associated with increased functional connectivity within the dorsal attention network, correlating to improvement in cognitive tests targeting attention | Ketones in MCI may be beneficial for cognition |
| Veneman et al., 1994 [60] | Immediate, ketone salt, healthy, no stressor | • **Primary Tests:** Cognitive Deterioration Test. | Infusion of BOHB increased the glycemic threshold and reduced magnitude of autonomic and neuroglycopenic symptoms and cognitive dysfunction | BOHB may substitute for glucose as a fuel for the brain and alter physiological responses in hypoglycemia |
| Waldman et al., 2018 [61] | Immediate, ketone salt, healthy, exercise | • **Primary Tests:** Cognitive Challenge (FitLights), Wingate Trials. | No significant differences among Wingate power output between treatments. | BHB did not improve high intensity cycling or cognitive performance measures |
| Waldman et al., 2019 [62] | Immediate, ketone salt, healthy, exercise | • **Primary Tests:** Dual-Stress Challenge, Mental Arithmetic Challenge, Stroop Color-Word | Blood BHB was elevated and remained throughout. Blood glucose was lower during KS | KS are not effective aids for enhancing cognitive performance during a DSC. |
|  |  | Test.<br>• **Secondary Tests:** Heart rate, Rating of Perceived Exertion (RPE), Blood BHB levels. | compared to PLA. No differences in HR, RPE, MAC, or SCW |  |
| Xu et al., 2020 [36] | Long-term, MCT, AD, no stressor | • **Primary Tests:** ADAS-Cog-C.<br>• **Secondary Tests:** Activities of Daily Living (ADL) scale. | ADAC-Cog-C scores were reduced for MCT intervention. ADL scores didn't change | MCT had positive effects on cognition in mild to moderate AD patients |
| Yomogida et al., 2021 [63] | Immediate, MCT, healthy, no stressor | • **Primary Tests:** Functional MRI (fMRI), Executive Function Tasks (N-back, Go/No-Go). | MCT meal improved N-back task | MCT was associated with heightened cognitive functions in healthy older adults. Potential beneficial impact of ketones with respect to improved cognitive outcomes. |

Many of the studies focused on long-term use of MCTs and structurally related EK compounds in older adults Several studies demonstrated improvements in cognitive performance in older adults with frailty, MCI or AD after long-term MCT supplementation [31–33], or coconut oil enriched Mediterranean diet [34]. EK were associated with increased MMSE scores and improvements in episodic memory, language, and executive function. Of note, two studies found that the cognitive benefits of long-term EK might be greater in individuals without the APOE4 allele [35, 36]. Only one long-term study utilized a non-MCT product, finding that 14 days of KME supplementation improved performance of the Digit Symbol Substitution Task in obese adults [8].

Of the studies investigating the immediate effects of EK supplements, a large proportion of these were in a younger, healthy population and utilized KME. The results were mixed, with some studies finding attenuation of exercise-induced decline in cognitive function [37] and visual reaction time [38] in the KME group, however, other studies used similar young, healthy populations in the context of exercise, or exercise plus hypoventilation found no protective effect of KME against cognitive decline [39, 40]. Two studies reported that a single MCT dose improved immediate cognitive task performance in AD [41] and in the context of hypoglycemia [42], although the effects were dose-dependent and varied by cognitive domain.

## Meta-analysis

Out of the 29 studies included in the systematic review, 18 were included in the meta-analysis, representing 875 participants, to quantify the effect of EK supplementation on cognitive function. The remaining 11 studies failed to report complete results (i.e., pre-post intervention), or presented the results as median.

First, we assessed the overall effect of EK on cognitive function, pooling results from 18 studies. The overall pooled effect was positive and statistically significant (SMD = 0.26, [95% CI 0.11 – 0.40], p = 0.0007) (using fixed effect model due to high heterogeneity (I² = 93%, p < 0.001) **Figure 3**). Risk of bias was assessed using funnel plot and Egger’s regression and did not show asymmetry (Egger’s intercept = 2.75 (0.87), p = 0.06, **Supplementary Figure 1**). The sensitivity analysis did not highlight the presence of outliers or studies with implausible results (extremely large effect) (**Supplementary Figure 2)**.

Next, we analyzed the differences between studies assessing the long-term (> 13 days) and immediate effects of EK. While no statistically significant difference between the duration groups (p = 0.50) was found, the overall effect was only statistically significantly different for studies assessing the long-term effect (SMD = 0.20 [0.11 – 0.28], p < 0.001, **Supplementary Figure 3**). For studies assessing the immediate effect, the results were at the border of the significance level (SMD = 0.31 [-0.01 – 0.64], p = 0.061).

When comparing the type of supplementation, the two most common supplement types, KME and MCT, were studied. As KME have a greater ketogenic effect than MCT [23, 43], we hypothesized that there might be a greater effect on cognition in KE studies. We did not find statistically significant differences between KME and MCT (p = 0.06, **Supplementary Figure 4**). However, the overall positive effect on cognition was only statistically significant for the MCT supplementation (SMD = 0.15 [0.05 – 0.25], p = 0.002) not for the KME (SMD = 0.31 [-0.07 – 0.68], p = 0.11). As only one study described the combination of MCT and BHB salt, it was not included in this analysis.

We then compared the cognitive effects of EK in studies of subjects with, and without the most included pathologies, MCI and AD. We expected that EK might provide greater benefit to subjects with a pathology that compromised their cognitive function. Surprisingly, we did not find a statistically significant difference (p = 0.21, **Supplementary Figure 5**) between the effect on healthy subjects (SMD = 0.32 [0.10 – 0.55], p = 0.004) and between subject with MCI or AD (SMD = 0.15 [0.01 – 0.30], p = 0.038).

We were interested to explore if the presence of a stressor such as exercise or hypoglycemia would modulate any effect of EK on cognition. However, no statistically significant differences (p = 0.25) were found between studies using no stressor (SMD = 0.17 [0.07 – 0.27], p < 0.001) and studies that included an exercise-based stress (SMD = 0.33 [-0.04 – 0.71], p = 0.081), although the latter was not significant in the standalone analysis.

Next, we performed meta-regression analysis to assess the relationship between cognitive outcomes and variables of interest. Firstly, we used meta-regression to determine if the duration of the EK intervention was related to cognitive outcomes. We did not find a significant relationship between the total duration of supplementation and cognitive outcomes for the overall dataset (β =-0.0007 (SE = 0.0013), p = 0.56, **Figure 4A**). However, when analyzed by subgroups, the data showed a tendency toward a positive effect of longer interventions for MCT supplementation, although this effect was not statistically significant (β = 0.001 (0.0006), p = 0.089), see **Table 3** for complete results.

**Table 3:**
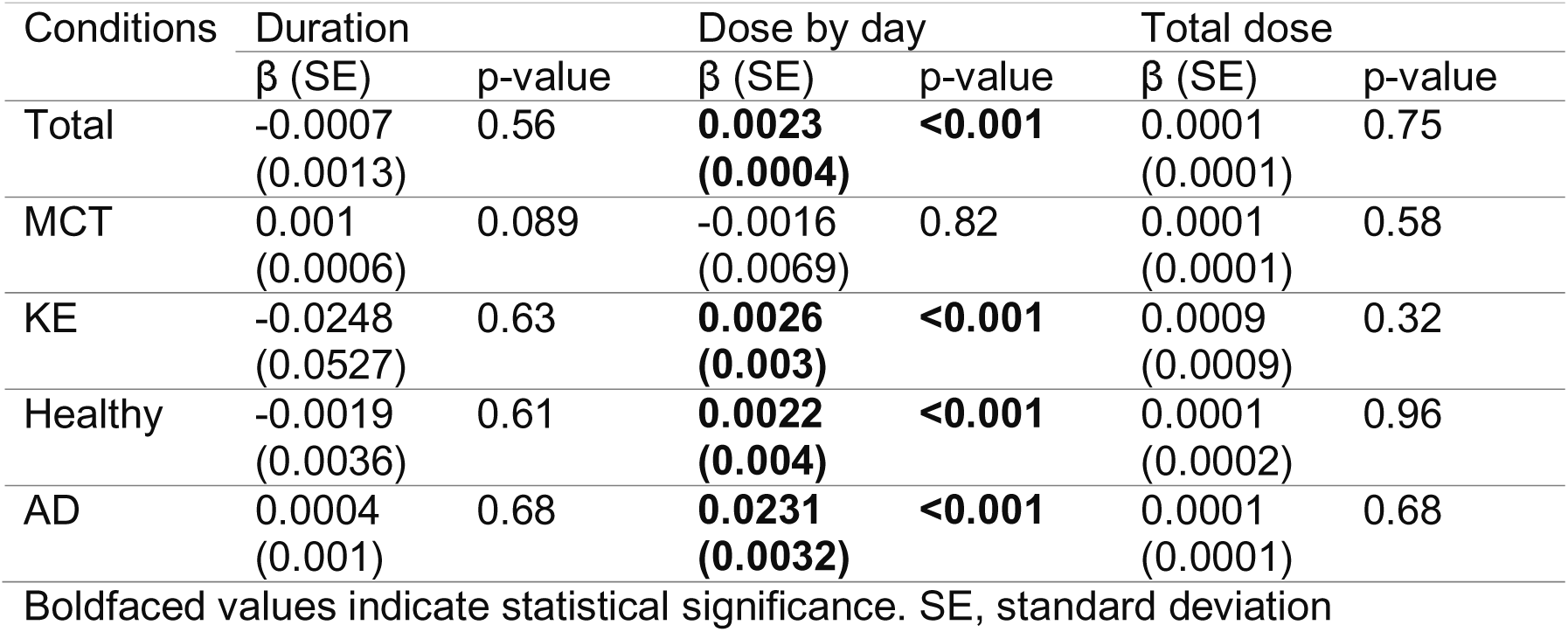
Meta-regression between the effect of ketone bodies supplementation on cognitive function for the total dose, the dose by day and the total duration of the supplementation.

| Conditions | Duration |  | Dose by day |  | Total dose |  |
| --- | --- | --- | --- | --- | --- | --- |
| | $\beta$ (SE) | p-value | $\beta$ (SE) | p-value | $\beta$ (SE) | p-value |
| Total | -0.0007<br>(0.0013) | 0.56 | **0.0023**<br>**(0.0004)** | **<0.001** | 0.0001<br>(0.0001) | 0.75 |
| MCT | 0.001<br>(0.0006) | 0.089 | -0.0016<br>(0.0069) | 0.82 | 0.0001<br>(0.0001) | 0.58 |
| KE | -0.0248<br>(0.0527) | 0.63 | **0.0026**<br>**(0.003)** | **<0.001** | 0.0009<br>(0.0009) | 0.32 |
| Healthy | -0.0019<br>(0.0036) | 0.61 | **0.0022**<br>**(0.004)** | **<0.001** | 0.0001<br>(0.0002) | 0.96 |
| AD | 0.0004<br>(0.001) | 0.68 | **0.0231**<br>**(0.0032)** | **<0.001** | 0.0001<br>(0.0001) | 0.68 |

Boldfaced values indicate statistical significance. SE, standard deviation

Secondly, we examined how the dose of EK consumed influenced the observed cognitive outcomes for the overall dataset. While a greater total EK dose over the study period did not significantly correlate with additional cognitive improvements (β = 0.0001 (0.0001), p = 0.75, **Figure 4D**), increasing daily dose of EK was strongly related to greater improvement in cognitive function across all studies (β = 0.0023 (0.0004), p < 0.001, **Figure 4C-D**). We then repeated the meta-regression to determine if the relationship between daily dose against and outcomes persisted in further subgroup analyses. The relationship between greater daily dose and cognitive improvements remained significant for studies using KME (β = 0.0026 (0.003), p < 0.001) and for both healthy participants (β = 0.0022 (0.0004), p < 0.001) and those with AD (β = 0.0231 (0.0032), p < 0.001). These findings imply that higher daily doses of EK consistently result in greater cognitive improvements across different populations and supplement types.

## Discussion

The aim of this systematic review and meta-analysis was to determine the effect of EK supplementation on cognitive function. The findings indicate that EK consumption can beneficially impact cognitive outcomes, with evidence highlighting the possible importance of both intervention duration and EK compound used and supporting efficacy in healthy and non-healthy adults. Overall, these findings highlight the promise of exogenous ketones as a novel strategy to target cognitive function, although further research is needed to support and inform clinical translation.

The positive effect of EK on cognition demonstrated by our overall, pooled analysis is in alignment with the primary teleological function of ketones as a back-up substrate for the brain during starvation [4]. However, considering the context of cerebral substrate provision during fasting (i.e., low carbohydrate availability) compared to in the studies included here (i.e., EK alongside sufficient carbohydrate) it is, in fact, perhaps surprising that cognitive function following EK exceeded function under conditions of normal, ample substrate availability. This observation hints at the importance of mechanisms beyond simple substrate oxidation to the functional effect of EK. A recent large observational study found a cognitive benefit of BHB concentrations over as low as ∼0.3 mM, at which oxidation would likely be low-to-minimal [28]. Recent clinical studies have proposed that EK may act to increase cerebral blood flow [7, 8], or increase brain network stability [44]. Preclinical data also demonstrates changes neurotransmitter release in the state of ketosis [10, 11]. Taken together, the additional cognitive benefit of EK in the context of sufficient carbohydrate substrate, suggests that ketones are not only a fuel for the brain, but that they have physiological signaling effects distinct from glucose [45] which translate to improved cognitive function.

The observation that only the ‘long-term’ (> 13 days) EK supplementation studies resulted in a significant effect on cognition in standalone analysis, but not ‘acute’ studies, suggests that consistent use of EK may be required for the emergence of a cognitive effect. However, our overall meta-regression did not highlight duration as a critical factor, with only a non-significant trend in the MCT subgroup analysis. Notably, there were very few long-term studies with KME. The possibility that EK efficacy builds over time is suggestive of a non-energy, signaling mechanistic contribution to their effect on cognition, as outlined above. If BHB’s role as an oxidative substrate were a primary driver of its effect on cognition, it would likely reach maximal effect while blood BHB was elevated as a substrate for cerebral metabolism. Ultimately, the latency of onset and persistence of cognitive effects following EK consumption is unknown; studies that help to define the latency and persistence will be foundational for any future translation of EK for cognitive function.

The results of our analysis leave many open questions about EK supplement type and dose selection. Whilst only studies using MCTs found a significant effect on cognition when analyzed as a standalone subgroup, the high heterogeneity in the KME studies made it difficult to detect an effect. The finding that daily dose was related to cognitive improvement in our meta regression is suggestive that higher, more sustained increases in blood ketones could be required for cognitive benefits. The only study to directly interrogate this was Fortier et al, who found that greater circulating ketone concentrations were associated with a greater cognitive improvement during a study of long-term MCT supplementation [32]. In other physiological systems greater ketone concentrations have been linked to larger functional changes, notably the cardiovascular system where cardiac output improves to a greater extent as ketone concentration increases [46]. As KME have a larger impact on circulating ketones concentrations that both MCT and ketone salts [23, 24], using these EK supplements in long term studies is a promising direction for future research. Despite their potent effect on blood ketone concentration, KME are known for a having a difficult to mask, bitter taste which could pose a barrier to long-term use. In contrast, both MCTs and medium chain fatty acid esters have a neutral taste, making them more translatable for long-term use.

There are several notable limitations to the generalizability of these results. The first, and the most important, is the high heterogeneity between studies. We noted significant variability in participant population, interventions, and outcome measures. This high level of heterogeneity (I² = 93%) across the studies, especially regarding the type of ketone supplementation and intervention duration, makes it difficult to generalize the findings. A second important limitation is that many of the studies were short-term interventions. There is a lack of long-term studies and long-term follow up, particularly with KME, which limits the understanding of the sustainability of the cognitive benefits. These limitations suggest that while the findings of the review are promising, more standardized, long-term studies with larger sample sizes are necessary to fully understand the efficacy of EK on cognition.

Despite these limitations, our results are promising and offer interesting perspective for future research and clinical translation. Given the beneficial cognitive effects of EK in both healthy and cognitively impaired individuals, no known safety concerns and acceptable tolerance profiles, EK are a candidate for further research and clinical translation. It is likely that the greatest effect would be seen if EK were used in synergy with other interventions, such as diet and exercise; this should be addressed in future research. No studies have addressed if EK can play a role in prevention, this too is an avenue of potential application. Other key questions highlighted by this analysis and foundational to the field of EK application at large include the choice of EK supplement type (MCT vs KME), dose selection and possible drivers of individual differences between individuals including metabolic responsiveness, cognitive and physical health.

In conclusion, this systematic review and meta-analysis found that exogenous ketone supplementation has a positive effect on cognitive performance. Future work should explore the relative contribution of ketones as fuel substrates vs signaling metabolite to the observed effects and determine if there is a threshold for dosing or blood ketone concentrations that delivers the greatest improvement with the lowest practical dosing.

## Abbreviations

AcAc: Acetoacetate
AD: Alzheimer’s Disease
ADAS-Cog: Alzheimer’s Disease Assessment Scale-Cognitive Subscale
ADRD: Alzheimer’s Disease and Related Dementias
BHB: D-β-hydroxybutyrate
EK: Exogenous ketones
kMCT: Ketogenic MCT
KME: Ketone monoester
MCI: Mild cognitive impairment
MCT: Medium chain triglyceride
mM: millimolar
MMSE: Mini mental state examination
PRISMA: Preferred Reporting Items for Systematic Reviews and Meta-Analyses
SD: Standard deviation of the mean

## Data Availability

All data produced in the present work are contained in the manuscript

